# Implications of delayed reopening in controlling the COVID-19 surge in Southern and West-Central USA

**DOI:** 10.1101/2020.12.01.20242172

**Authors:** Raj Dandekar, Emma Wang, George Barbastathis, Chris Rackauckas

## Abstract

In the wake of the rapid surge in the Covid-19 infected cases seen in Southern and West-Central USA in the period of June-July 2020, there is an urgent need to develop robust, data-driven models to quantify the effect which early reopening had on the infected case count increase. In particular, it is imperative to address the question: How many infected cases could have been prevented, had the worst affected states not reopened early? To address this question, we have developed a novel Covid-19 model by augmenting the classical SIR epidemiological model with a neural network module. The model decomposes the contribution of quarantine strength to the infection timeseries, allowing us to quantify the role of quarantine control and the associated reopening policies in the US states which showed a major surge in infections. We show that the upsurge in the infected cases seen in these states is strongly co-related with a drop in the quarantine/lockdown strength diagnosed by our model. Further, our results demonstrate that in the event of a stricter lockdown without early reopening, the number of active infected cases recorded on 14 July could have been reduced by more than 40% in all states considered, with the actual number of infections reduced being more than 100, 000 for the states of Florida and Texas. As we continue our fight against Covid-19, our proposed model can be used as a valuable asset to simulate the effect of several reopening strategies on the infected count evolution; for any region under consideration.

## 2 BACKGROUND

The Coronavirus respiratory disease 2019 originating from the virus “SARS-CoV-2”^1, 2^ has led to a global pandemic, leading to more than 50 million confirmed global cases in more than 200 countries as of November 13, 2020.^3^ In the United States, the first infections were detected in Washington State as early as January 20, 2020,^4^ and now it is being reported that the virus had been circulating undetected in New York City as early as mid-February.^5^ As of September 21, 2020, the United States has ≈ 6.9 million infected cases since the virus began to spread.

Since the second week of June, a second surge of Covid-19 was seen in the United States,^6^ with rapidly increasing daily infected cases, hospitalization rates and death rates.^7, 8^ Initially driven by disastrous situations in the states of Arizona, South Carolina, Texas, Florida and Georgia,^6^ the surge in cases was also later seen in several other Southern and West-Central states.^9^ This surge can be seen in figure 1 which shows the active infected cases over time as of July 14, 2020 with a 7-day moving average for 9 states. States which reopened early show a generally strong co-relation with the rise in the infected cases over the 3-month period from late April to mid July 2020.^9^ For example, states which opened before May 15 showed daily infected case increments of: Florida (1393 %), Arizona (858 %), South Carolina (999 %), Alabama (547 %), Oklahoma (477 %), Tennessee (279 %), Georgia (245 %), Mississippi (215 %), Nevada (697 %), Texas (680 %) and Utah (287 %); while states which reopened after May 29 showed values of: Michigan (16 %), Pennsylvania (–26 %), New York (−52 %), New Jersey (−32 %) and Illinois (− 54 %). Thus, although early reopening seems to be co-related to the second surge of cases seen in the USA, there is a need for robust, data-driven quantification of the effect of early reopening on the growth of infected count data. More importantly, it is of utmost importance to answer the question: How many infected cases could have been prevented, had the worst affected states not reopened early?

**Figure 1:**
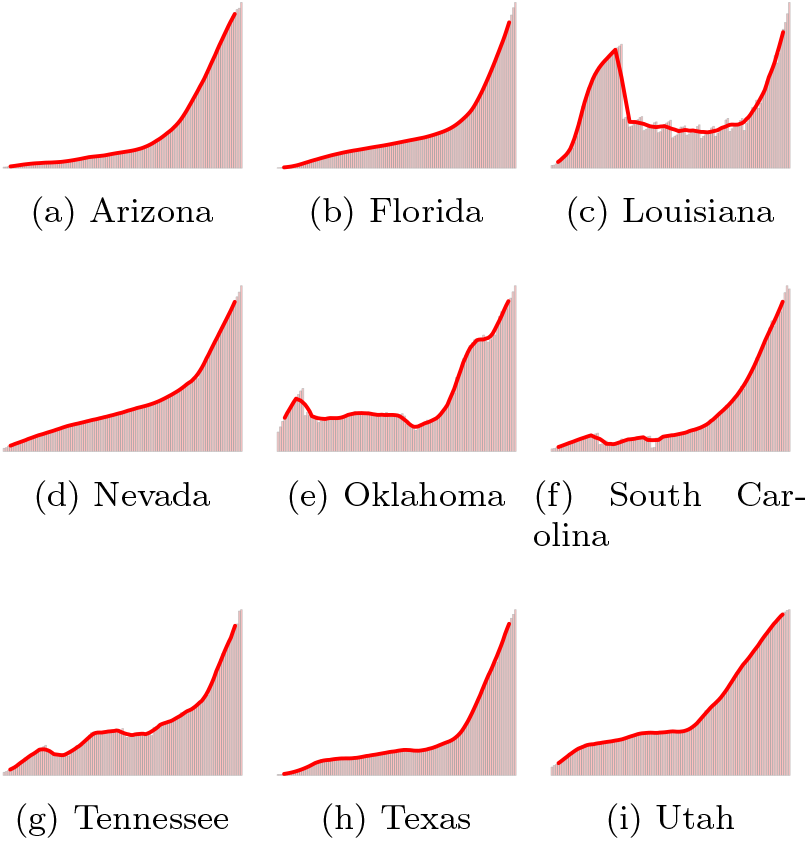
Active infected cases over time as of July 14, 2020, shown with a 7-day moving average, for the Southern and West-Central states considered in the present study.

In an effort to address this question, we have developed a machine learning-aided epidemiological model. The novelty of our model arises from the fact that it allows us to decompose the contribution of quarantine/lockdown strength evolution to the infected data timeseries for the region under consideration. This enables us to simulate the effect of varying quarantine strength evolutions and hence varying reopening strategies on the infected count data. We define reopening as beginning when a state allows its stay-at-home order to expire, or, in the case of states that never issued a stay-at-home order, when a state first starts allowing non-essential businesses, such as dine-in restaurants and hair salons, to reopen.^10, 11^ The reopening details for the states considered in the study are shown in table 1. Considering nine US states which showed a significant surge in cases since the last month, we demonstrate that our model shows a drop in the quarantine strength evolution when these states were reopened. Furthermore, we show that maintaining a strict lockdown without early reopening would have led to about 500, 000 fewer infected cases in all these states combined.

**Table 1:** Reopening details for different states considered in the present study

|  |  |  |
| --- | --- | --- |
| State | Reopening date | Reopening details |
| 1. Arizona | May 15 | June 17: Mask regulations strengthened,<br>June 29: Partial reversal of reopening |
| 2. Florida | May 4 | June 3: Phase 2 of reopening |
| 3. Louisiana | May 15 | June 5: Phase 2 of reopening |
| 4. Nevada | May 9 | May 26: Phase 2 of reopening |
| 5. Oklahoma | April 24 | May 15: Phase 2 of reopening,<br>June 1: Phase 3 of reopening |
| 6. South Carolina | May 4 | May 4: Stay at home order lifted,<br>further facilities reopened till May 18 |
| 7. Tennessee | April 30 | May 22: Phase 2 of reopening. |
| 8. Texas | May 1 | May 18: Phase 2 of reopening,<br>June 3: Phase 3 of reopening |
| 9. Utah | May 1 | May 1: Gradual reopening |

## 3 METHODS

### 3.1 QSIR Model

#### Standard SIR model

The SIR (Susceptible - Infected - Recovered) is governed by the following set of ODEs

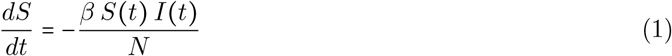

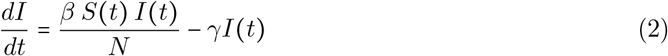

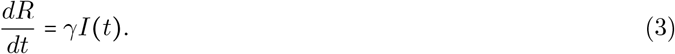

where *β, γ* are the contact and recovery rates respectively. We use this framework as our baseline model to be augmented with an neural network module. We do not consider the possibility of recovered individuals being reinfected.^12^ We also do not consider the waning of immunity associated with Covid-19 as discovered in recent studies.^13^

#### QSIR model: ODE formulation

The QSIR ODE model formulation is similar to the one studied previously,^14^ and is briefly explained in this section. The equations governing the QSIR model are as follows

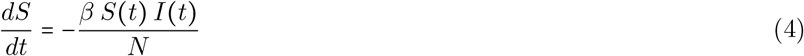

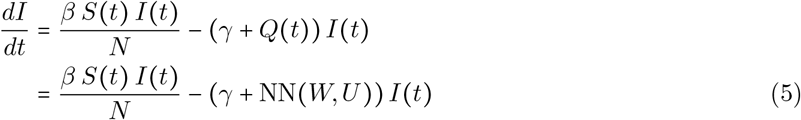

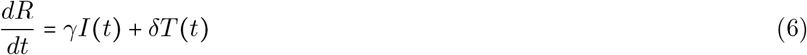

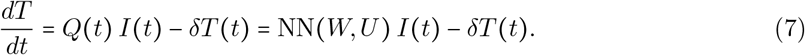

The SIR model is augmented by introducing a time varying quarantine strength rate term *Q*(*t*) represented by a neural network^15^ and a quarantined population *T*(*t*), which is prevented from having any further contact with the susceptible population. Thus, the term *I*(*t*) denotes the active infected population (Actively infected = Cumulative infected - Recovered) still having contact with the susceptibles, as done in the standard SIR model, while the term *T*(*t*) denotes the infected population who are effectively quarantined and isolated.

#### Augmented QSIR Model: Initial Conditions

The starting point *t* = 0 for each simulation was the day at which 500 infected cases was crossed, *i*.*e. I*_0_ ≈ 500. The number of susceptible individuals was assumed to be equal to the population of the considered region. Also, in all simulations, the number of recovered individuals was initialized from data at *t* = 0 as defined above. The quarantined population *T*(*t*) is initialized to a small number *T*(*t* = 0) ≈ 10.

### Augmented QSIR Model: Parameter estimation

The data for the infected, recovered case counts was obtained from the publicly maintained repository by the Center for Systems Science and Engineering at John Hopkins University. The loss function is defined as

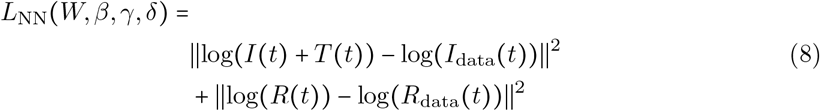

Parameter optimization for *W, β, γ, δ* was performed by minimizing the loss function defined in Equation 8 using the approach employed in prior studies^14, 16, 17^ using an ADAM optimizer^18^ with a learning rate of 0.01. For most of the states under consideration, *W, β, γ, δ* were optimized by minimizing the loss function given in (8). For states with a low recovered count: Arizona, Florida, Nevada and Texas, we employed a two stage optimization procedure to find the optimal *W, β, γ, δ*. In the first stage, (8) was minimized. For the second stage, we fix the optimal *γ, δ* found in the first stage to optimize for the remaining parameters: *W, β* based on the loss function defined just on the infected count as *L*(*W, β*) = ∥log(*I*(*t*) + *T*(*t*) −log*I*_data_(*t*))∥^2^. Such an approach was found to be optimal for analyzing low recovered count data in previous studies.^14^

In all states considered in the present study, we trained the model using data starting from the dates when the 500^th^ infection was recorded in each region and up to July 14, 2020. For each state considered, *Q*(*t*) denotes the rate at which infected persons are effectively quarantined and isolated from the remaining population, and thus gives composite information about (a) the effective testing rate of the infected population as the disease progresses and (b) the intensity of the enforced quarantine as a function of time.

This QSIR ODE framework applied on the infected and recovered data is used to estimate the quarantine strength function Q(t) in a particular state as shown in the first and second columns of figure 3.

**Figure 2:**
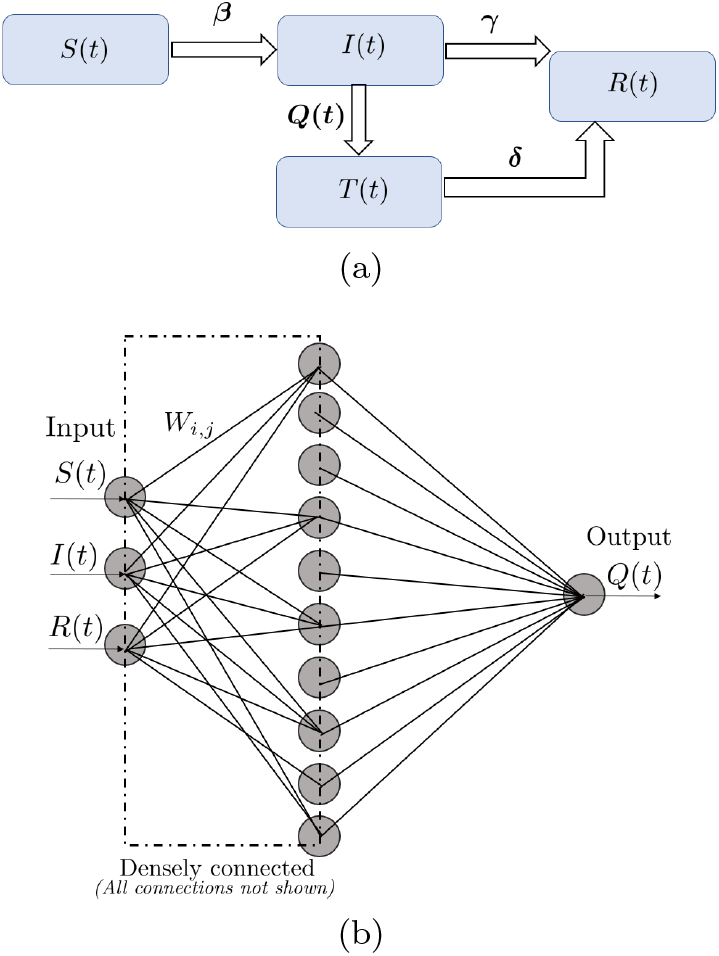
(a) Schematic of the augmented QSIR model considered in the present study. (b) Schematic of the neural network architecture used to learn the quarantine strength function *Q*(*t*).

**Figure 3:**
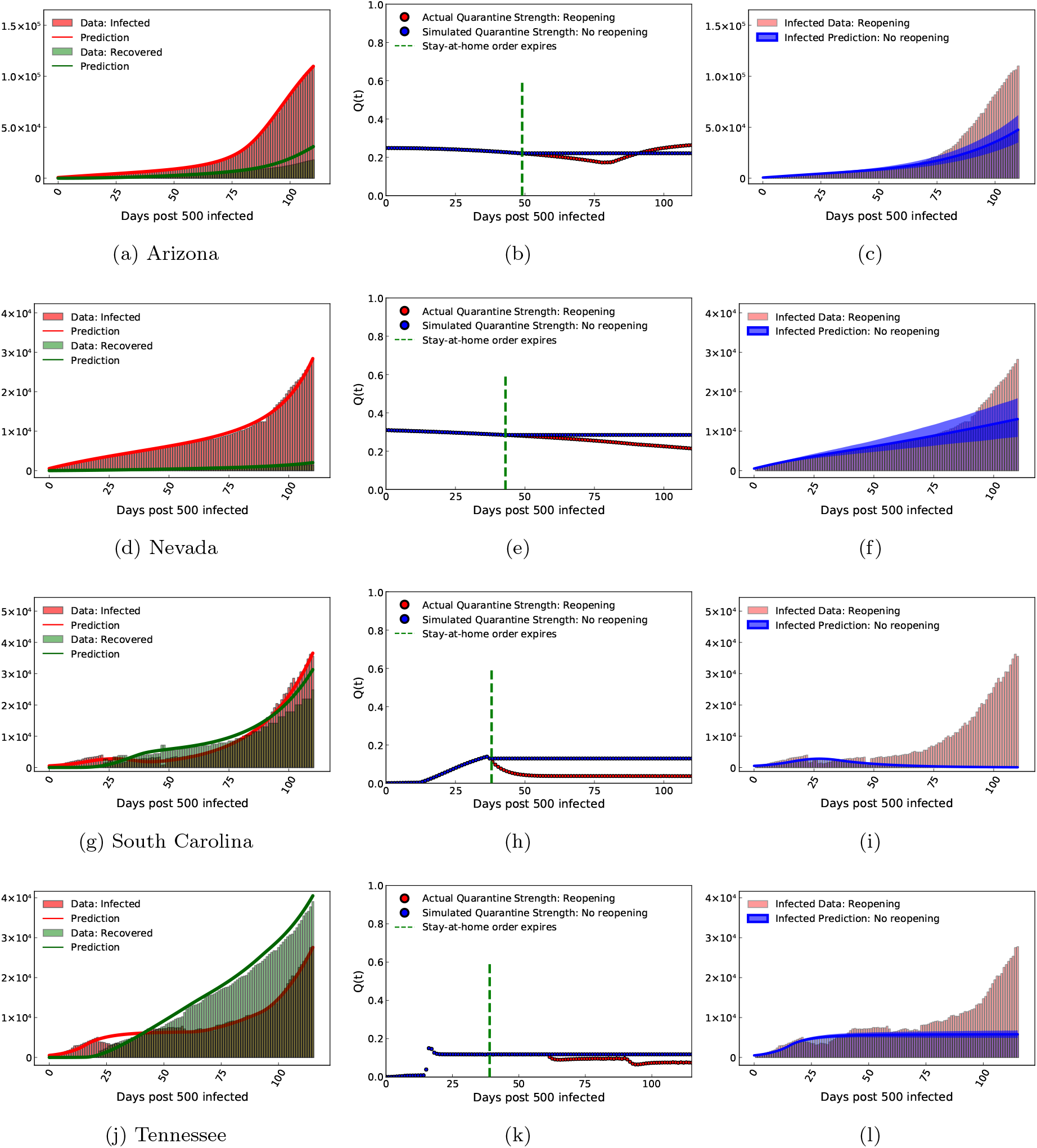
For the states of Arizona, Nevada, South Carolina and Tennessee, figure shows: (a, d, g, j) Model recovery of infected and recovered case count as of 14 July, 2020. (b, e, h, k) Quarantine strength function as discovered by our trained model (with reopening). This is shown along with the quarantine strength function which we use to simulate strict quarantine without reopening after stay-at-home order was imposed. (c, f, i, l) Estimated infected count if strict quarantine and lockdown measures were followed without reopening (5% and 95% quantiles are shown) as compared to the values corresponding to the actual early reopening scenario.

### QSIR Model: SDE formulation

The ODE modelling framework described above is a deterministic approach to model transfer of species (here: people) from one compartment to another through different reaction channels. Such a deterministic approach ignores any random fluctuations during species transfer from one compartment to the other. To include such stochastic effects and thus get a measure of the model uncertainty, we note that the augmented SIR framework derives from the chemical master equation which descibes the time evolution of the probability of such a system of interacting species to be in a given state at a given time (details in Supplementary Information). Although the chemical master equation cannot be solved analytically, under certain conditions, it can be distilled down to a stochastic differential equation (SDE) which captures the fluctuations in species transfer as random walks. Such an SDE, also known as the Chemical Langevin Equation, is thus based on the underlying ODE framework (macroscopic picture) and also includes stochastic effects reminiscent of microscopic modelling. In fact, in the Supplementary Information, we show that the microscopic simulation, macroscopic ODE formulation and the Chemical Langevin Equation (which acts as a bridge between the two) are all equivalent to each other.

The equivalent stochastic formulation or the Chemical Langevin equation for the augmented SIR model is

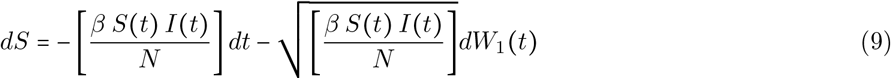

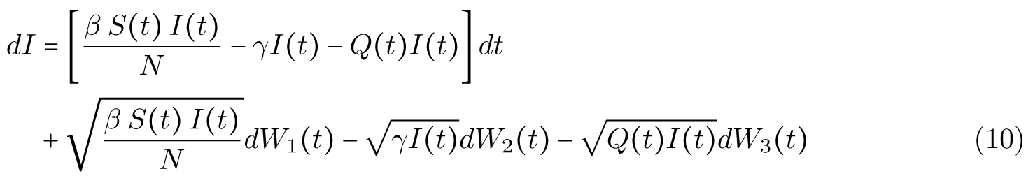

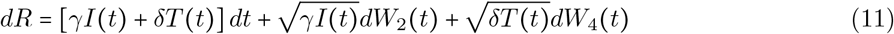

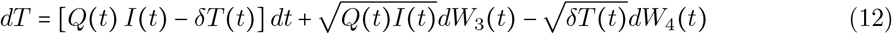

In (9), *W*_*i*_ (*t*) ∼ *N*(0, *t*) is a normally distributed random variable with mean zero and variance *t* or *dW*_*i*_(*t*) ∼ *N*(0, *dt*). It should also be noted that each *W*_*i*_(*t*) represents an independent Brownian motion. The simulations were performed using the Catalyst.jl software in Julia using the LambaEM algorithm based on.^19^ 1000 trajectories were simulated for each state.

This QSIR SDE framework along with the simulated quarantine functions for no reopening is used to predict the new infected case count and hence estimate the reduction in the number of infected cases under the simulated no-reopening quarantine function. The results are shown as 5% and 95% quantiles in the third column of figure 3.

### Mean Absolute Percentage Error

The Mean Absolute Percentage Error (MAPE) is defined as

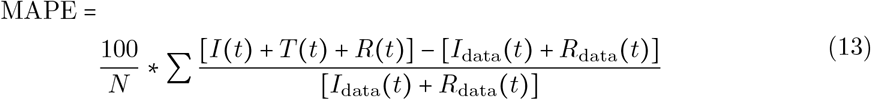

where *N* is the number of observations.

## 4 RESULTS

The first stage of our analysis is using our model,^14^ called the QSIR model to diagnose the underlying quarantine strength evolution *Q*(*t*) in the regions under consideration. By applying the QSIR model to more than 70 countries globally, we have established the validity of *Q*(*t*) in accurately diagnosing the on-the-ground quarantine situation in majorly affected European, South American and Asian countries.^14^ A slow growth of *Q*(*t*) without a significant increase indicates relaxed quarantine policies, a sharp transition point in *Q*(*t*) is indicative of a sudden ramp-up of quarantine measures, and an inflection point corresponds to the time when the quarantine response was the most rapid in the region under consideration. The results of our model applied globally to all continents are hosted publicly at covid19ml.org.

In this study, to perform the quarantine diagnosis to analyze the implications of delayed reopening, we applied the QSIR model to 9 US states which showed a significant surge in the infected case count in the last month: Arizona, Florida, Louisiana, Nevada, Oklahoma, South Carolina, Tennessee, Texas and Utah. Figure 3 shows representative results for Arizona, Nevada, South Carolina and Tennessee. The plots for the remaining states are provided in the Supplementary Information. Figures 3 a, d, g, j show the comparison of the infected and recovered count estimated by our model with the actual data. A reasonable agreement is seen for all states, with the model being able to capture the rise in infections seen in the tail end of the timeseries. The QSIR model details are provided in the Methods section; Mean Absolute Percentage Error (MAPE) values for the model along with the epochs required for convergence for each state are provided in Supplementary Information.

Figures 3 b, e, h, k show the quarantine strength evolution *Q*(*t*) as learnt by the neural network module, which shows a decline whose starting point corresponds well to the time when these states began reopening, as seen from table 1 and the green dotted line in the figures 3 b, e, h, k. In some states, the decline in *Q*(*t*) starts later than the reopening date; possibly corresponding to the Phase 2 or Phase 3 of reopening (table 1) or because of the time delay for population level changes to be seen in the infected count evolution, after reopening. *Q*(*t*) trained by our model shows a significant drop after early reopening in all Southern and West-Central states that showed a surge in cases last month; whereas the North-Eastern states of New York, New Jersey and Illinois, which reopened late and showed no surge in infections, did not show a drop in *Q*(*t*) (Table 2 and figures in Supplementary Information). Thus, the upsurge in the infected cases seen in these states is strongly co-related with a drop in the quarantine/lockdown strength *Q*(*t*) diagnosed by our model. This is indicative of two things: (a) the Southern and West-Central states reopened early, which led to a relaxed imposition of quarantine/lockdown measures in these states and consequently a surge in infections was seen, and (b) the North-Eastern states of New York, New Jersey and Illinois reopened late, and even after reopening, a relatively low contact rate was maintained amongst the population, leading to a relatively high magnitude of the imposed quarantine strength, which prevented a surge of infections in these states.

**Table 2:** Drop in quarantine strength function, *Q*(*t*) after reopening as discovered by our trained model. *Q*(*t*) trained by our model shows a significant drop for all Southern and West-Central states which showed a surge in cases from reopening; whereas the North-Eastern states which showed no surge don’t see a drop in *Q*(*t*).

| State | Reopening date | % increase in daily cases<br>since reopening | Maximum % decrease<br>in $Q(t)$ after reopening |
| --- | --- | --- | --- |
| 1. Arizona | May 15 | +858 | +22 |
| 2. Florida | May 4 | +1393 | +10 |
| 3. Louisiana | May 15 | +193 | +30 |
| 4. Nevada | May 9 | +697 | +25 |
| 5. Oklahoma | April 24 | +477 | +29 |
| 6. South Carolina | May 4 | +999 | +71 |
| 7. Tennessee | April 30 | +279 | +44 |
| 8. Texas | May 1 | +680 | +29 |
| 9. Utah | May 1 | +287 | +39 |
| 10. New York | May 29 | -52 | -45 |
| 11. New Jersey | June 9 | -32 | -60 |
| 12. Illinois | May 29 | -54 | -8 |

After confirming that our model is able to accurately depict the co-relation between the surge in infections and early reopening in these states through the diagnosed *Q*(*t*), we proceed to the second in stage of our analysis. In the second stage, we use the diagnosed *Q*(*t*) to address the question: How many infected cases would have been reduced, had the worst affected states not reopened early? To answer this question, we simulate the “no-reopening” strategy by assuming that *Q*(*t*) is maintained at the value it was before reopening, without decreasing. This simulated *Q*(*t*) is shown in Figures 3 b, e, h, k. The flexibility of our model allows us to run our model with this simulated *Q*(*t*) for all states considered. To quantify the aleatory uncertainty resulting from random fluctuations in the model, we utilized the chemical Langevin equation extension to the QSIR model whose definition and justification is described in the Methods and Supplemental Information section. This allows us to estimate bootstrapped confidence intervals resulting from 1000 simulations of such a stochastic model, and thus quantify the effect of such a “no-reopening policy” on the epidemic spread. The infected count evolution for the simulated *Q*(*t*) without reopening is shown in Figures 3 c, f, i, l (5% and 95% quantiles are shown). We can see that, for all these states, instead of seeing a spike in infections, we would have seen a plateau in the infected case count evolution. The number and the percentage of infected cases that would have been prevented by July 14 had these states not reopened are shown in Table 3. It is evident that the number of infections could have been reduced by more than 40% in all states considered, with the actual number of infections reduced being more than 100, 000 for the states of Florida and Texas. Even the less populated states of Louisiana, South Carolina and Tennessee show mean infected case reduction values of 44%, 84% and 47% respectively, which correspond to 36, 000, 51, 000, and 31, 000 infected cases reduced.

**Table 3:** Infected count reduction by 14 July, 2020, if states had not reopened early, as estimated by our model.

| State | % decrease<br>(5% - 95% quantiles) | Mean<br>% decrease | Case reduction | Mean case<br>case reduction |
| --- | --- | --- | --- | --- |
| 1. Arizona | 35 - 62 | 49 | 44000 - 79000 | 63000 |
| 2. Florida | 20 - 75 | 49 | 57000 - 218000 | 144000 |
| 3. Louisiana | 37 - 50 | 44 | 31000 - 41000 | 36000 |
| 4. Nevada | 32 - 68 | 51 | 10000 - 20000 | 15000 |
| 5. Oklahoma | 46 - 69 | 58 | 10000 - 15000 | 13000 |
| 6. South Carolina | 83 - 86 | 84 | 50000 - 52000 | 51000 |
| 7. Tennessee | 41 - 53 | 47 | 27000 - 36000 | 31000 |
| 8. Texas | 41 - 51 | 46 | 115000 - 143000 | 129000 |
| 9. Utah | 35 - 47 | 41 | 11000 - 14000 | 12000 |

## 5 CONCLUSION

In this study, we have developed a novel methodology to quantify the effect of early reopening on the infected case count surge seen during the period of June-July 2020. We have proposed a machine learning model, called the QSIR model, rooted firmly in fundamental epidemiology principles which has the following attributes: (a) it is highly interpretable with few free parameters rooted in an epidemiological model, (b) it relies on only Covid-19 data and not on previous epidemics and (c) it can decompose the infected timeseries data to reveal the quarantine strength/policy variation, *Q*(*t*), in the region under consideration. To demonstrate the validity of our model in capturing the actual quarantine policy evolution in a particular region, the model has been applied to 70 countries globally. The quarantine strength behaviour learnt from the model accurately mimics the on-the-ground situation in majorly affected European, South American and Asian continents. The results for this global analysis are hosted at covid19ml.org.^14^

After confirming our belief in the model through a global analysis, we apply the model to the Southern and West-Central US states which have shown a massive surge in Covid-19 infected cases since June 2020. We demonstrate that the *Q*(*t*) extracted by our model shows a significant drop in value for the Southern and West-Central states which reopened early and showed a surge in infections. The time at which *Q*(*t*) starts to decline generally agrees well with the reopening date for the states considered. Since the decline in *Q*(*t*) is strongly co-related to the surge of infections and also the reopening date for states which reopened early, we can then simulate the effect of “no-reopening” by maintaining the *Q*(*t*) at a constant level after reopening, instead of declining. We show that maintaining a steady imposition of quarantine/lockdown control would have played a massive role in bringing down the infected count by more than 40% in all states considered, with the infections reduced reaching more than 100, 000 for the states of Florida and Texas.

We have proposed a novel machine learning methodology, rooted in fundamental epidemiological models; which is able to recover the real time quarantine strength evolution for any region under consideration. As the pandemic evolves and we continue our fight against Covid-19; and for future outbreaks, our globally applicable methodology can be a valuable asset for researchers and policy makers to simulate several reopening strategies, counterfactual scenarios and analyze their impact on the infected count evolution. Our findings highlight that as we continue the fight against Covid-19, it is imperative to reduce the contact between susceptible and infected individuals in public places by formulating robust safety guidelines. Such guidelines implemented and maintained in the affected states would ensure a high level of quarantine strength associated with that state and can prevent a future surge or wave in the Covid-19 infected count timeseries.

Validation of the model robustness and parameter identifiability have been mentioned in the Supplementary Information. We have also compared an equivalent of the effective reproduction number called the Covid spread parameter in our study, with other studies to further validate the results of our modelling approach. The Covid spread parameter is defined by (a) the infected individuals and (b) the recovered individuals from both the infected and the quarantined states; since both of those effectively don’t further contribute to the infection spread.^14^

The results of our model should be taken in the context of its assumptions. Ideally, one needs to consider the shifting US testing policies for the time period under consideration. Since the testing efforts did not show a significant increase during and after the reopening in the US states in the time period considered within the present study^20, 21^ and we did not want to burden our model with additional parameters to fit; testing compartments have not been included in the present study. Additionally, several studies in literature^22–25^ have attempted to incorporate underreporting of infected/recovered cases in their modelling paradigm. Most of these studies use previously known estimates of testing data, serology data or Infection-Fatality-Rate(IFR). In these studies involving multiple parameters, a number of parameters are assumed to fixed at the start of the simulation from prior studies. These parameters include and are not limited to: time between onset of infections and symptoms, transmission duration, rate at which hospitalized patients recover,^25^ mean duration from symptom onset to recovery^22^ or even the IFR ratio.^22^ A second class of studies uses antibody testing from collected serum samples to estimate the actual number of infected cases.^26^

As the pandemic unfolds and starts spreading, the first information available is the number of infected, recovered and deaths (for example: the John Hopkins public repository for Covid-19 tracking). Unless we have serum sample data information or we can confidently rely on prior studies for assessment of certain parameters, accurate information of the underreporting factor is difficult to obtain in real time. One of the goals of the present modelling methodology is to assist researchers and policy makers with quarantine diagnosis information in real time, with no reliance on parameters derived from prior studies.

Finally, the model is based on the SIR framework, which assumes a constant, age-independent contact and recovery rate between the infected and susceptible populations. Additionally, we do not consider the spatial heterogeneity in the infected count within a particular state and assume the governing dynamics to be only time-dependent. Consideration of these second-order aspects would further refine the model and would be the subject of future studies.

## 6 Supplementary Information

### Model-diagnosed quarantine strength for North-Eastern US states

Figure 4 shows the application of the model to the north-eastern states of New York, New Jersey and Illinois along with the diagnosed quarantine strength function *Q*(*t*) for these states. These states do not show a decline in *Q*(*t*). This corresponds well to the delayed reopening and generally stronger quarantine measures employed in the North-Eastern US states. Since *Q*(*t*) does not decrease, these states did not show a surge in infections starting June 2020, unlike their Southern and West-Central counterparts. The difference in these results between the North-Eastern and Southern, West-Central states indicates two things: (a) it strengthens the validity of our proposed model in capturing the real-time reopening scenario in different states through the evolution of the diagnosed *Q*(*t*), and, more importantly, (b) it further validates the role played by early reopening in reducing *Q*(*t*) and subsequently leading to a surge of new infected cases in the Southern and West-Central US states.

**Figure 4:**
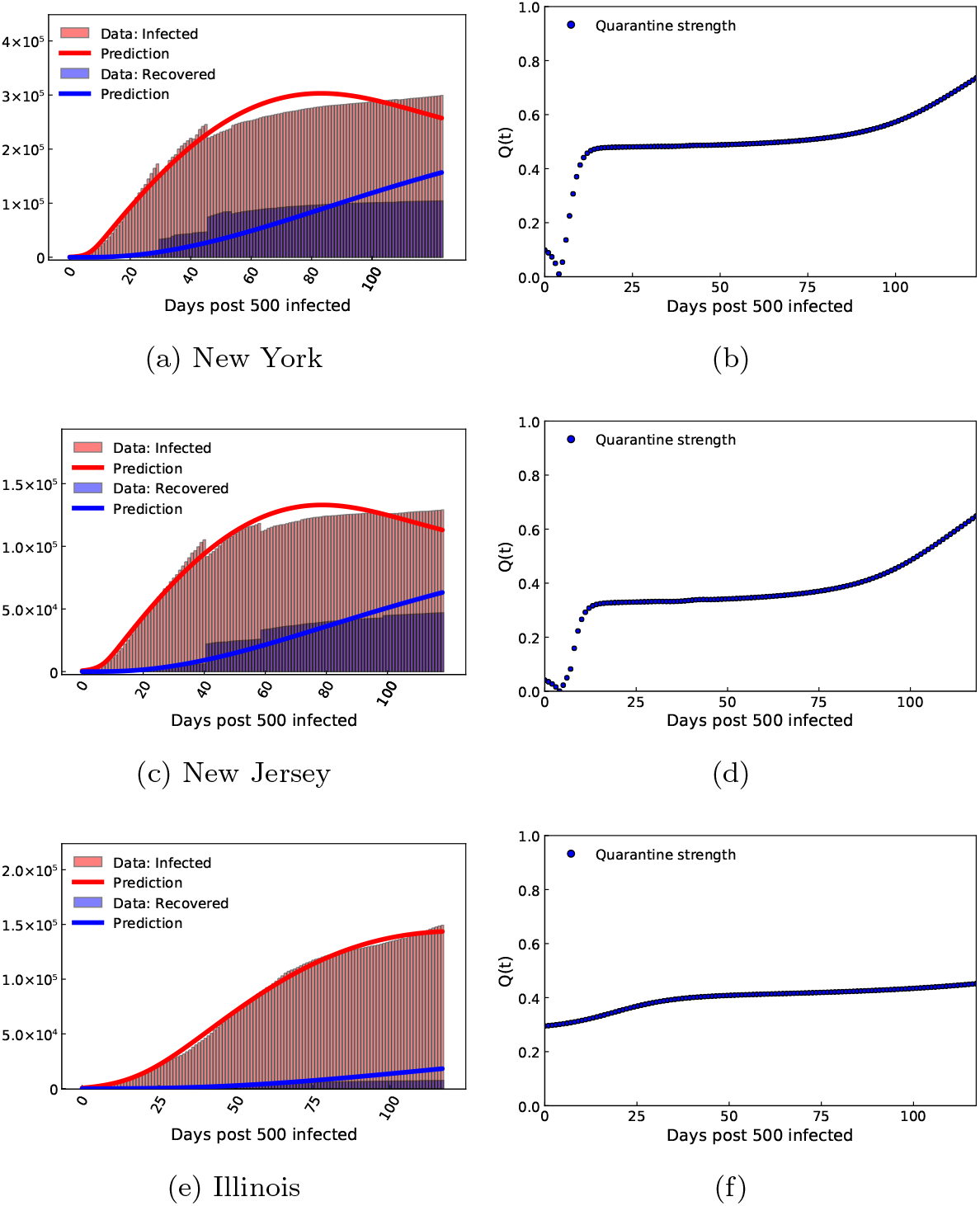
For the states of New York, New Jersey and Illinois, figure shows: (a, c, e) Model recovery of infected and recovered case count trained until 14 July, 2020. (b, d, f) Quarantine strength function as discovered by our trained model

### Impact of early reopening on the states of Louisiana, Florida, Oklahoma, Texas and Utah

Figure 5, 6 implements a similar analysis to study the effect of early reopening for the states of Louisiana, Nevada, Oklahoma, Texas and Utah, as done for the states of Arizona, Nevada, South Carolina and Tennessee. Similar to the states considered in the main text, we see that all of these states show a decline in *Q*(*t*) starting around the time when these states were reopened. If these states were not reopened early, a large number of infections would have been reduced as demonstrated in Table 1 of the main text.

**Figure 5:**
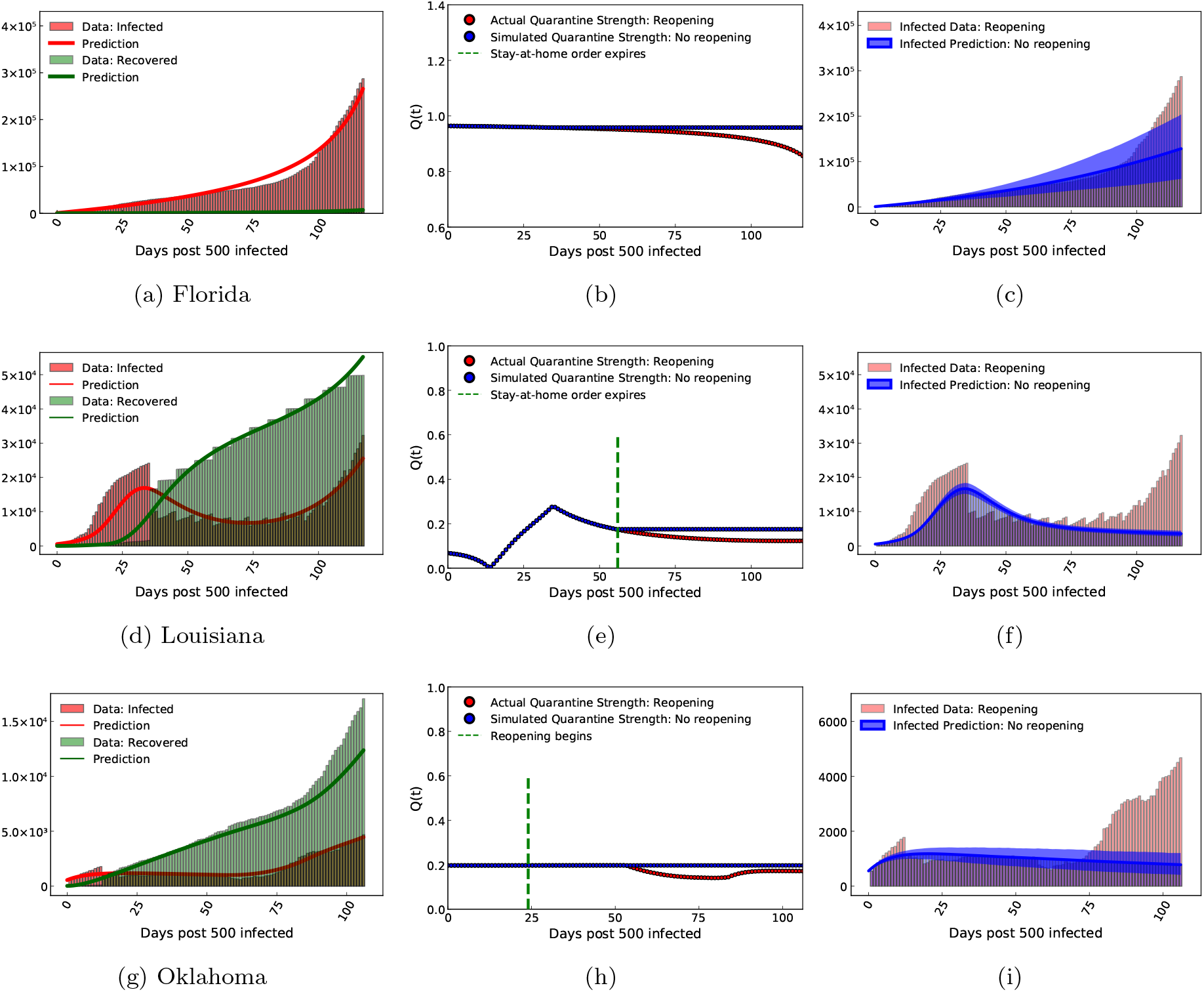
For the states of Louisiana, Nevada and Oklahoma: (a, d, g) Model recovery of infected and recovered case count as of 14 July, 2020. (b, e, h) Quarantine strength function as discovered by our trained model (with reopening). This is shown along with the quarantine strength function which we use to simulate strict quarantine without reopening after stay-at-home order was imposed. (c, f, i) Estimated infected count if strict quarantine and lockdown measures were followed without reopening as compared to the values corresponding to the actual early reopening scenario.

**Figure 6:**
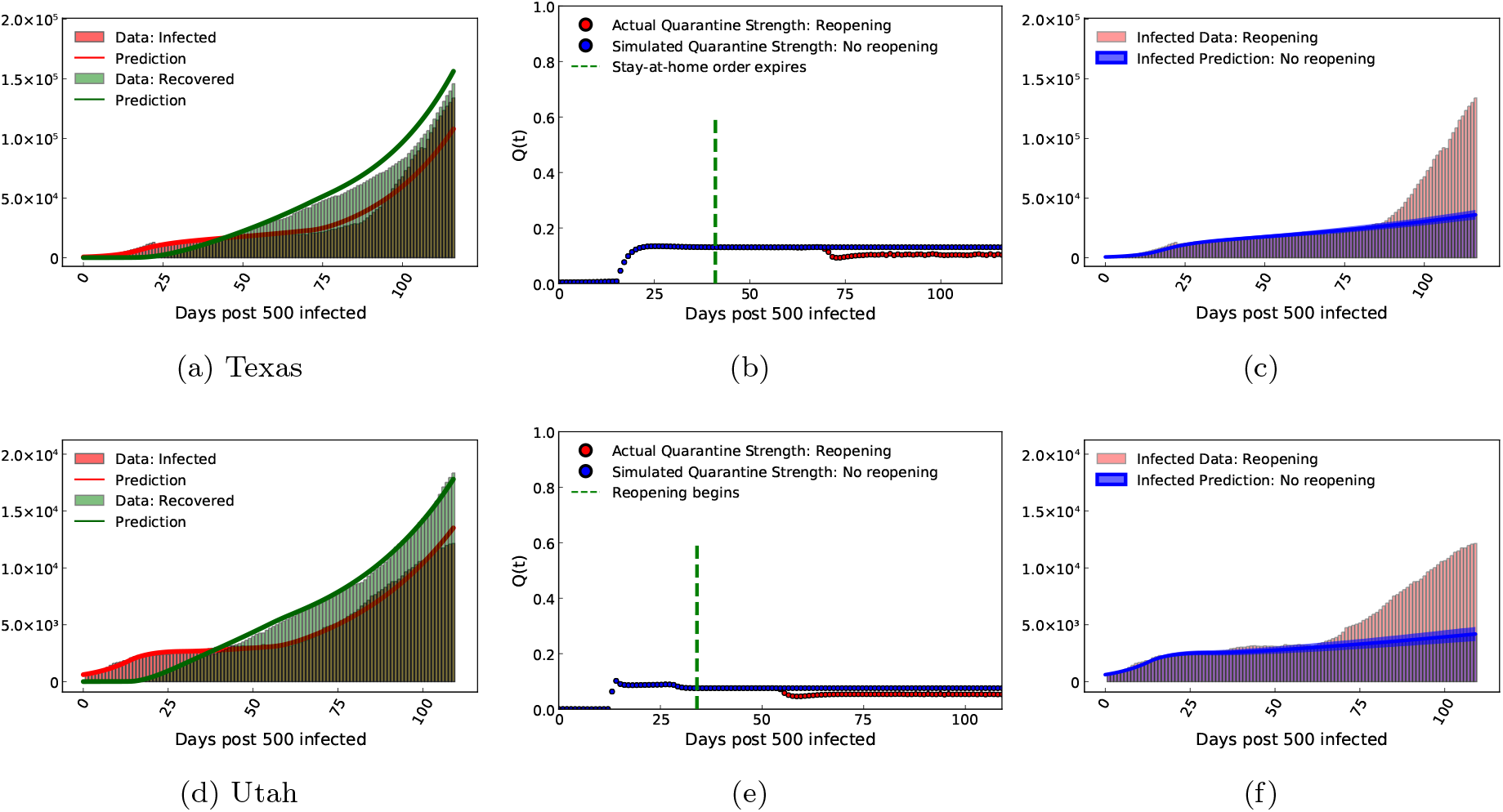
For the states of Texas and Utah: (a, d) Model recovery of infected and recovered case count as of 14 July, 2020. (b, e) Quarantine strength function as discovered by our trained model (with reopening). This is shown along with the quarantine strength function which we use to simulate strict quarantine without reopening after stay-at-home order was imposed. (c, f) Estimated infected count if strict quarantine and lockdown measures were followed without reopening as compared to the values corresponding to the actual early reopening scenario.

### Equivalence between the ODE model and the Chemical Langevin SDE model

This analysis heavily borrows from the pioneering work done by Gillespie.^27^ In this section, we will establish that the deterministic ODE model and the stochastic Chemical Langevin equation originate from a common expression: the chemical master equation,^28^ and are closely linked to one another. Following is the notation we will use, in accordance with^27^ We consider *N* compartments: *S*_1_, *S*_2_ … *S*_*N*_ and *R* reaction channels: *R*_1_, *R*_2_ … *R*_*M*_ in a fixed volume Ω. In our case, we have *N*= 4 (*S, I, R, T*) compartments and *R* = 4 reaction channels. We denote the dynamical state of the system at any time *t* as *X*(*t*) = (*X* _1_ (*t*), *X*_2_ (*t*) … *X*_*N*_ (*t*)) where

- *X*_*i*_ (*t*): total number of *S*_*i*_ molecules (in our case: individuals) in the system.
- Propensity function *a*_*j*_(*x*)*dt* : probability that a reaction *R*_*j*_ will occur somewhere in Ω in the next time interval [t, t+dt] for *j* = 1, 2 … *M*.
- State change vector *ν*_*j*_ whose *i*th component is defined by *ν*_*j,i*_: change in the number of *S*_*i*_ molecules produced by one *R*_*j*_ reaction for *i* = 1, 2 … *N, j* = 1, 2 … *M*. In our case *ν*_*j,i*_ = ±1

From the definition of *a*_*j*_ (*x*)*dt*, we can write the probability of the system being in state *x* at time *t* + *dt* (we take the sum of all mutually exclusive ways either through one reaction or no reaction in [t, t+dt]):

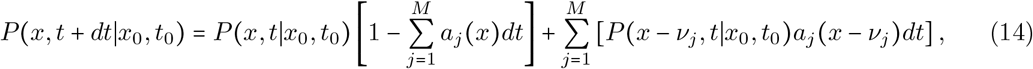

Taking the limit of (14) as dt -¿ 0 leads to the **chemical master equation**

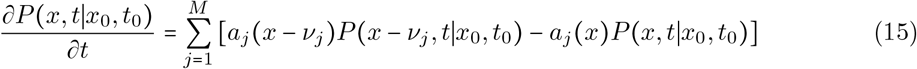

#### Macroscopic picture: Deterministic model relation to the chemical master equation

Multiplying the chemical master equation (15) by *x*_*i*_ and summing over all *x*, we obtain for the mean of *X*_*i*_ (*t*)

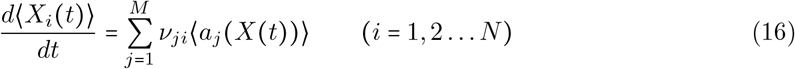

Thus, whenever fluctuations are not important, the species populations evolve deterministically according to the following set of ordinary differential equations

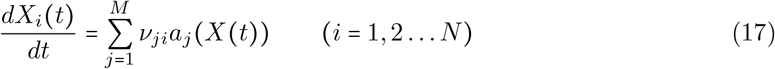

(17) is the basis for the classical SIR epidemiological equations, and we see how they evolve from the chemical master equation (15).

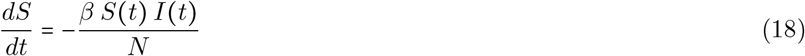

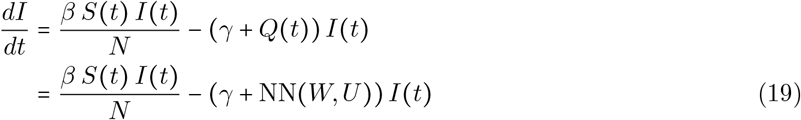

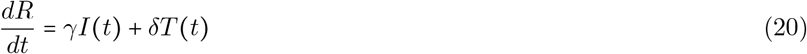

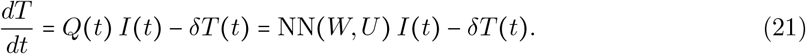

The ODE system used in the present study shown in (5-8), is of the form (17).

#### Microscopic picture: Stochastic Simulation Algorithm and its relation to the master equation

Another consequence of the master equation (15) is the existence and form of the next-reaction density function *p*(*τ, j*|*x, t*), which is defined as

- *p*(*τ, j*|*x, t*)*dτ* = probability that given *X*(*t*) = *x*, the next reaction in Ω will occur in [*t* + *τ, t* + *τ* + *dτ*], and will be an *R*_*j*_ reaction

Since ∑_*j*_ *a*_*j*_(*x*)*dt* is the probability that some reaction occurs in the time interval *dt*, the probability that a time interval *τ* is spent without any reaction occuring is given by the exponential distribution: Exp(∑_*j*_ *a*_*j*_ (*x*) *τ*). Thus, we obtain for *p*(*τ, j*|*x, t*)

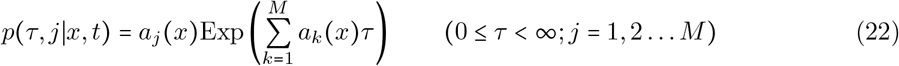

(22) is the basis for the stochastic simulation algorithm in which Monte-Carlo techniques are used to construct unbiased realizations of the process *X*(*t*). A typical algorithm for stochastic simulation of this kind, is the Gillespie Algorithm^29^ which can be viewed as a discrete space continuous time Markov jump process, with exponentially distributed jump times.

#### Chemical Langevin Equation: Bridging the gap between macroscopic and microscopic models

Let the state of the system *X*(*t*) at the current time *t* be *x*_*t*_. Let *K*_*j*_ (*x*_*t*_, *τ*) be the number of *R*_*j*_ reactions that occur in the time interval [t, t+dt]. Thus, the number of *S*_*i*_ molecules in the system at time *t* +*τ* will be

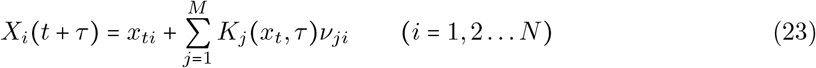

^27^ approximated *K*_*j*_ by imposing the following conditions

- **Condition 1: No propensity function change** This condition requires *τ* to be small enough so that none of the propensity functions *a*_*j*_ (*x*) change noticeably. The propensity functions then satisfy

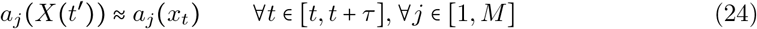 Due to this condition, *K*_*j*_(*x*_*t*_, *τ*) will be a statistically independent Poisson random variable *P*_*j*_ (*a*_*j*_ (*x*_*t*_), *τ*). Thus (23) simplifies to

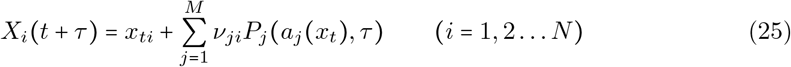
- **Condition 2: Large number of reaction occurrences:** This condition requires *τ* to be large enough so that the expected number of occurrences of each reaction channel *R*_*j*_ in [*t, t* +*τ*] is much larger than 1. Thus

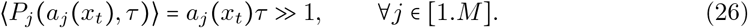 This condition enables us to approximate each Poisson variable *P*_*j*_ (*a*_*j*_ (*x*_*t*_), *τ*) by a normal random variable with the same mean and variance. Thus, (25) further simplifies to

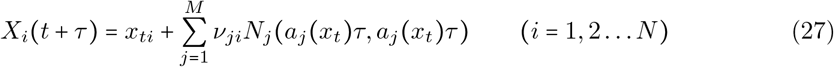

where *N*(*m, σ*^2^) denotes the normal random variable with mean *m* and variance *σ*^2^. Using *N*(*m, σ*^2^) = *m* + *σN*(0, 1), denoting the time interval *τ* by *dt* and the unit normal random variable *N*_*j*_ (0, 1) as *N*_*j*_ (*t*), we obtain

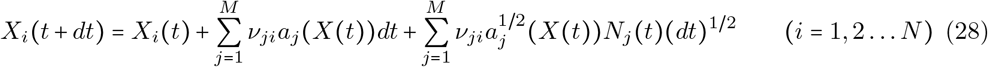 (28) can be written as a stochastic differential equation as

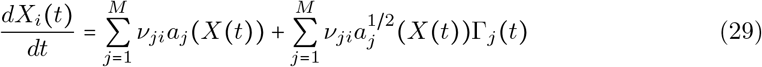

where Γ_*j*_ (*t*) are temporally uncorrelated, statistically independent Gaussian white noise processes. (29) is the Langevin equation, and it derives from the master equation provided that Condition 1 and Condition 2 are satisfied.

The Langevin equation (29) form of the ODE system (5-8) leads to the stochastic differential equation used in the current study

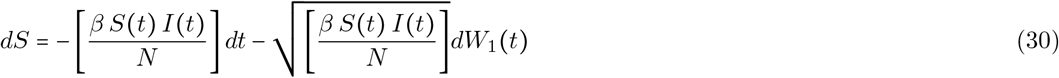

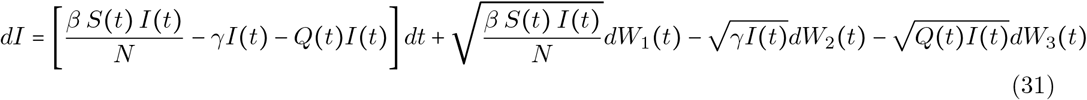

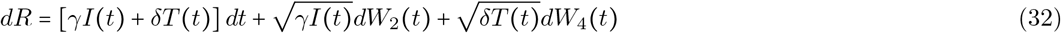

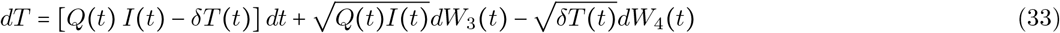

In (30), *W*_*i*_(*t*) ∼ *N*(0, *t*) is a normally distributed random variable with mean zero and variance *t* or *dW*_*i*_(*t*) ∼ *N*(0, *dt*). It should also be noted that each *W*_*i*_(*t*) represents an independent Brownian motion.

#### Comparison of the macroscopic, microscopic and Langevin SDE model for our study

Figure 7a shows that the microscopic Stochastic Simulation Gillespie Algorithm and the ODE model presented in Equation (6-9) in the main text show a good agreement with each other. Figure 7b shows the comparison of the Chemical Langevin SDE model shown in (30) ran for 1000 trajectories and the ODE model; which also show a good agreement. Thus, we have shown the equivalence between the microscopic, macroscopic and the Chemical Langevin model for our study. This equivalence allows us to add fluctuating components to the standard deterministic SIR model as shown in (30) and quantify the uncertainty resulting from these fluctuations.

**Figure 7:**
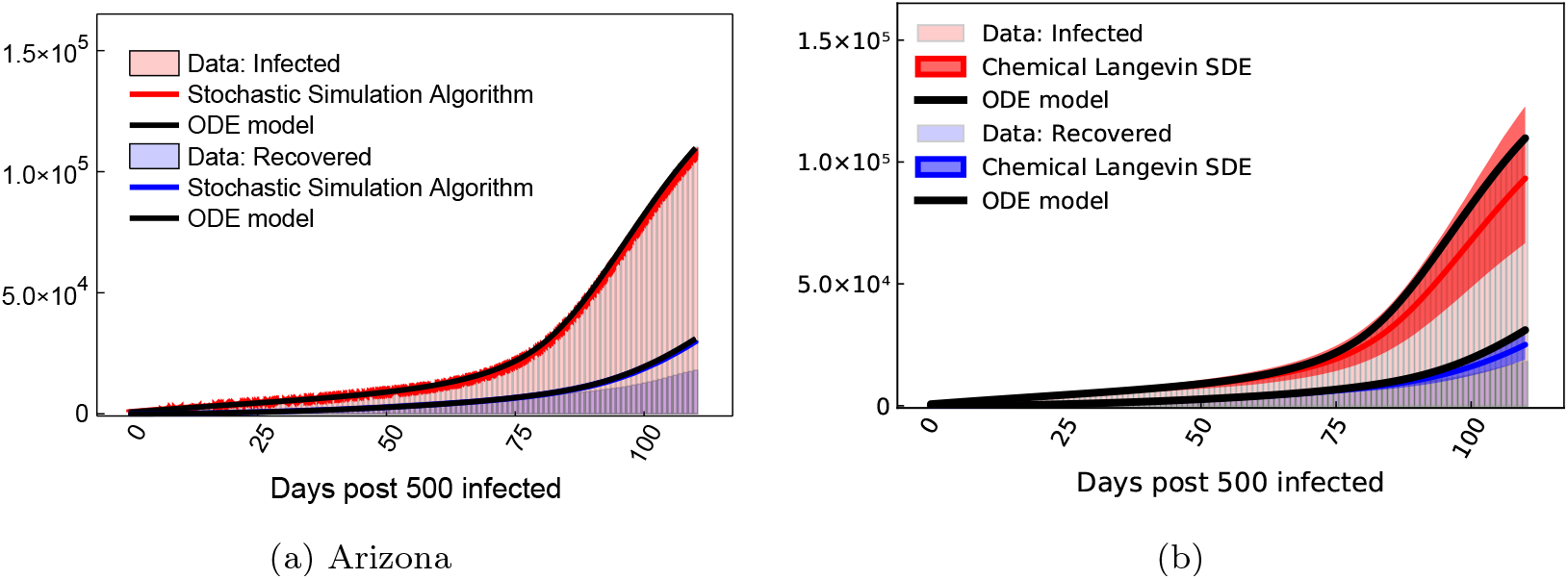
(a) Comparison of the microscopic Stochastic Simulation Gillespie Algorithm and the ODE model presented in Equation (6-9) in the main text. (b) Comparison of the Chemical Langevin SDE model shown in (30) ran for 1000 trajectories (5%*and*95% quantiles are shown) and the ODE model.

### Model specifications for each state

Table 4 shows the Model Mean Absolute Percentage Error (MAPE), epochs needed for convergence and number of parameters optimized for the different states considered.

**Table 4:** Mean Absolute Percentage Error (MAPE) values are shown along with the number of epochs required for and the number of parameters optimized, for all states considered.

| State | Model MAPE | Epochs | Parameters optimized |
| --- | --- | --- | --- |
| 1. Arizona | 5.4% | $10^5$ | 54 |
| 2. Florida | 18.7% | $10^5$ | 54 |
| 3. Louisiana | 12% | $12^5$ | 54 |
| 4. Nevada | 3.14% | $18^5$ | 54 |
| 5. Oklahoma | 7.9% | $12^5$ | 54 |
| 6. South Carolina | 11.7% | $12^5$ | 54 |
| 7. Tennessee | 6.9% | $12^5$ | 54 |
| 8. Texas | 10.4% | $24^5$ | 54 |
| 9. Utah | 3.79% | $12^5$ | 54 |

### Parameter Inference: Gaussian Process Residue Model

In order to validate the robustness of the model and the uniqueness of the parameters recovered by the model, we consider a Gaussian Process residue model for uncertainty quantification. Gaussian Processes have emerged as a useful tool for regression, classification, clustering and uncertainty quantification.^30, 31^

In the present study, we fit a Gaussian Process regression model between the error resulting from the best fit model and the infected data. For the prior over the function space, we use a mean of zero and variance described by a Squared Exponential Kernel with a lengthscale of 1 and a significantly high signal standard deviation of *O*(10^4^) which allows for noisy estimates of the posterior. Such a fitted model for the infected count for a region under consideration (Arizona), is shown below in figure 8. Subsequently, we sample 500 error residues from this model and superimpose them on the best fit predictions to simulate 500 samples of the infected case count data. Finally, we apply our model described on these 500 samples of data, and recover the parameters *Q* (*t*), *β, γ, δ* from each of them.

**Figure 8:**
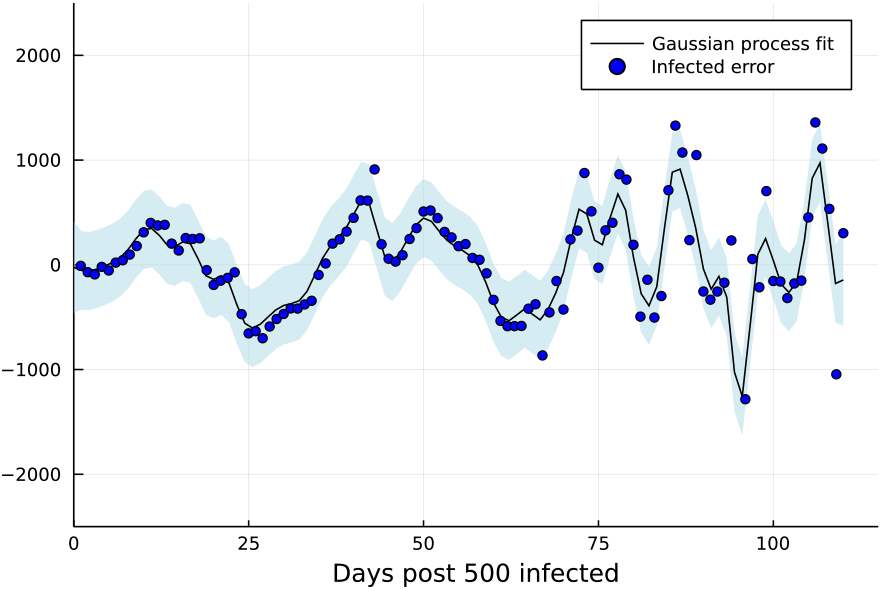
[Gaussian Process Residue Regression Model] Gaussian Process residue model fitted to the infected case count shown for Arizona.

Figures 9, 10 shows the inferred parameters for 500 realizations of the Gaussian process residue model superimposed on the best fit model prediction applied to all states considered, and shown for (a) the quarantine strength function *Q*(*t*), (b) the contact rate *β* and (c) the recovery rate *γ* + *δ*. It can be seen that for all realizations, *Q*(*t*) is seen to follow a similar behaviour, which lies close to the best fit model prediction. In addition, the inferred histograms for the contact rate *β* and the recovery rate *γ* +*δ* show a peak which is close to the best fit model prediction. This further validates the robustness of the model for other regions considered and strengthens the uniqueness of the parameters recovered by the model. A total of 12 million iterations (60000 iterations for each realization of the Gaussian process residue model × 500 realizations) were performed on the MIT Supercloud cluster to generate parameter histograms for each state considered.

**Figure 9:**
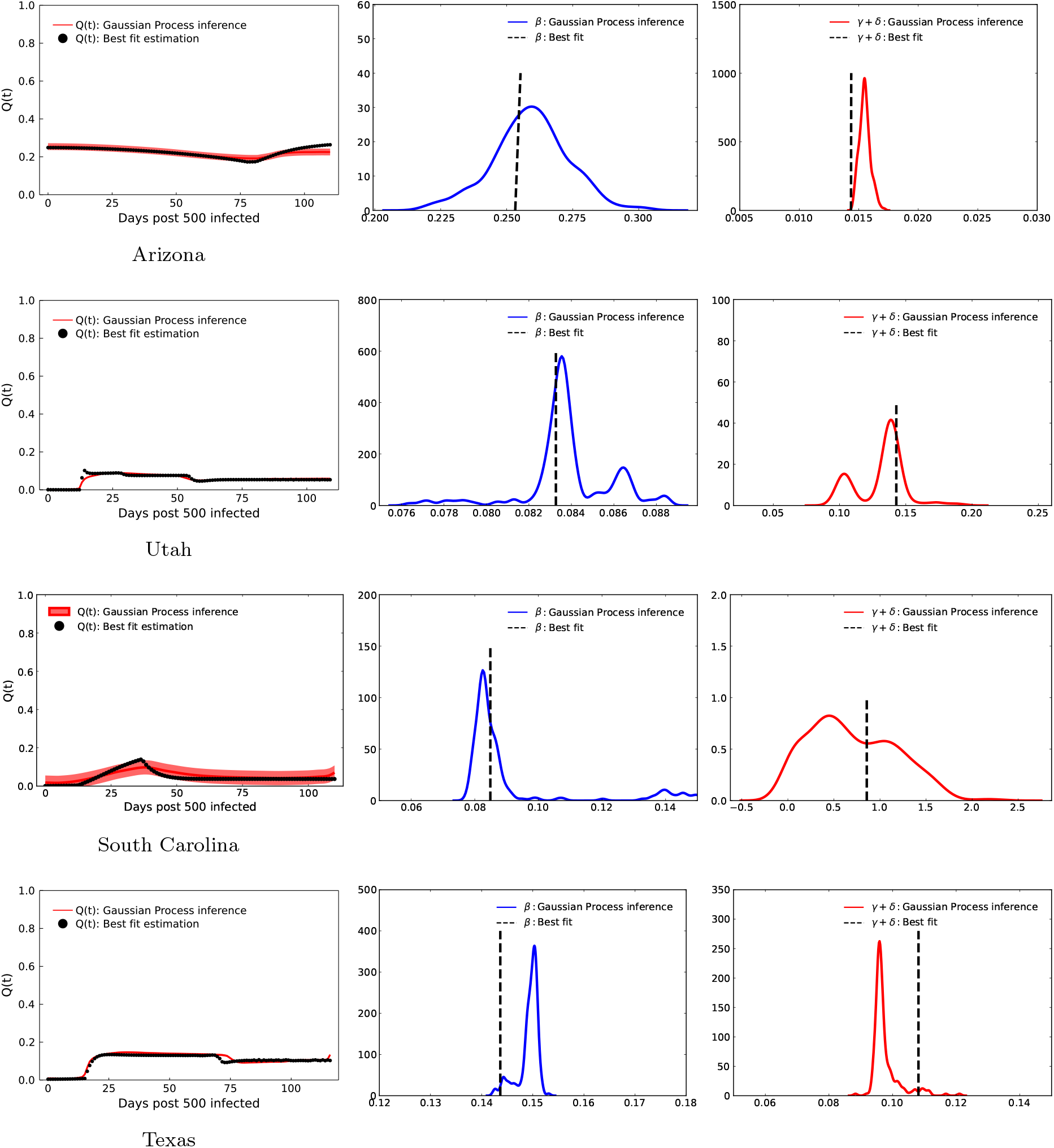
[Parameter Inference for US states] Inferred parameters for 500 realizations of the Gaussian process residue model superimposed on the best fit model prediction applied to the region considered for demonstration, and shown for (a) the quarantine strength function *Q*(*t*), (b) the contact rate *β* and the recovery rate *γ* + *δ*. A total of 12 million iterations were performed on the MIT Supercloud cluster to generate parameter histograms for one state.

**Figure 10:**
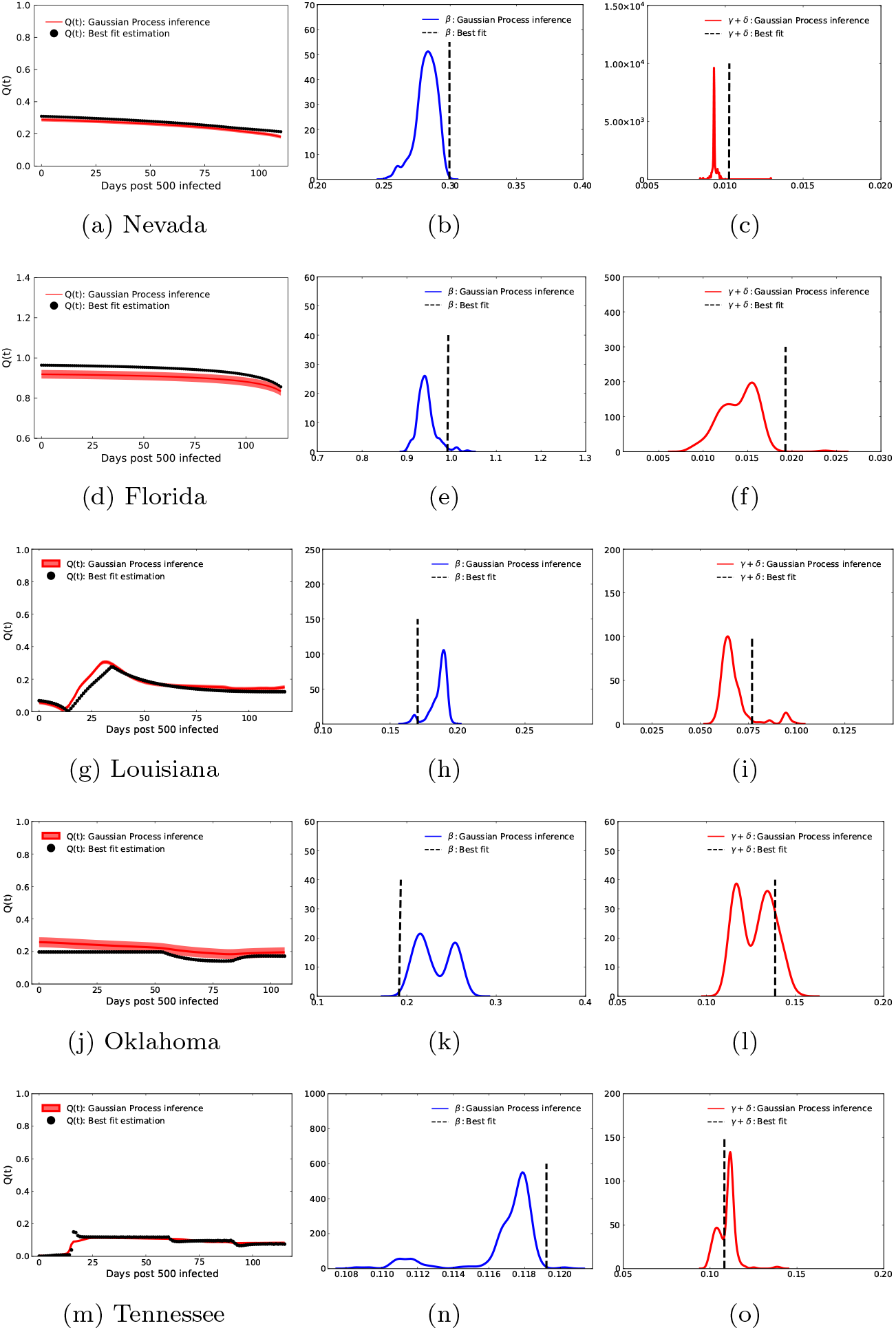
[Parameter Inference for US states] Inferred parameters for 500 realizations of the Gaussian process residue model superimposed on the best fit model prediction and shown for the quarantine strength function *Q*(*t*) (left column), the contact rate *β* (middle column) and the recovery rate *γ* +*δ* (right column) for the US states considered in the present study. A total of 12 million iterations were performed on the MIT Supercloud cluster to generate parameter histograms for each region.

### Model validation: Calculation of the effective reproduction number

Following a previous study,^14^ we define the Covid spread parameter as follows:

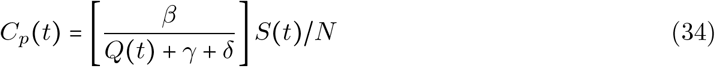

where *S*(*t*) is the susceptible population and *N* is the total population. This definition of the Covid spread parameter *C*_*p*_(*t*) is equivalent to the effective reproduction number *R*_*eff*_(*t*) in the context of the QSIR model. We included both *γ, δ* in the definition of *C*_*p*_(*t*) since both these parameters eventually contribute to the recovered population and we wanted to include effects of both. Another viable option to define *C*_*p*_(*t*) could be to just use *γ* in the denominator of *C*_*p*_(*t*).

Figure 11 shows the comparison of the Covid spread parameter, as defined in Equation 34 with and without reopening for all US states considered in the present study. For all the states, we can see that without reopening, a diminished effective reproduction number is seen, indicating moving in the right direction of halting the infection spread.

**Figure 11:**
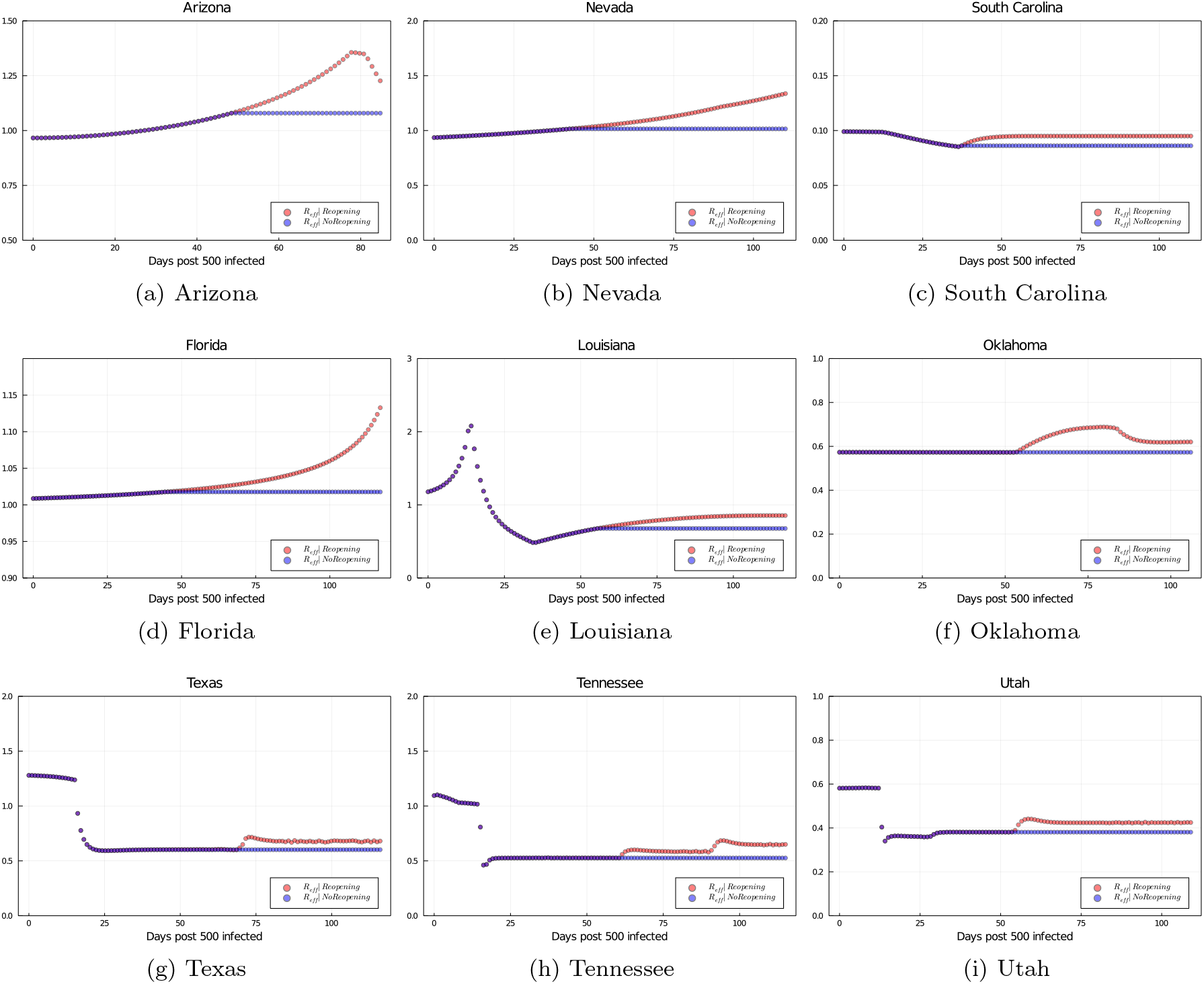
The comparison of the effective reproduction number, as defined in Equation 34 with and without reopening, shown for all US states considered in the present study

To further validate the Covid spread parameter variation and its relation to the effective reproduction number, we compare the variation in *C*_*p*_ to the *R*_*eff*_ obtained through a prominent Covid-19 forecasting model used by the CDC, USA.^20, 21^ For all of the 9 states which we considered, the time at which an upsurge is seen in *C*_*p*_ due to early reopening corresponds very well to the exact time at which an upsurge is seen in *R*_*eff*_ .^20, 21^ In addition, we show the comparison between *C*_*p*_ values estimated from our study and *R*_*eff*_ values obtained from^20, 21^ from reopening till one month post that; in table 5. For the states of Arizona, Nevada, Louisiana, Florida, Texas and Tennessee, these values lie close to each other. This further validates the results of our study and the quantitative metrics derived therein.

**Table 5:** *C*_*p*_ and *R*_*eff*_ value ranges from reopening till one month post that, for 6 states considered in our study; lie close to each other.

| Region | $C_p$ range<br>(Our study) | $R_{eff}$ range<br>(Ref.<sup>20</sup>) |
| --- | --- | --- |
| Arizona | 1.1 – 1.4 | 1.15 – 1.3 |
| Florida | 1 – 1.1 | 1.07 – 1.64 |
| Nevada | 1.05 – 1.5 | 1.19 – 1.5 |
| Louisiana | 0.7 – 1 | 0.88 – 1.62 |
| Texas | 0.6 – 0.7 | 1.08 – 1, 3 |
| Tennessee | 0.5 – 0.65 | 0.97 – 1.07 |

## Data Availability

All code files and results are publicly available at https://github.com/RajDandekar/Reopening_ImpactSimulator_US_States

## 7 DATA AND CODE AVAILABILITY

Data for the infected and recovered case count in all regions was obtained from the Center for Systems Science and Engineering (CSSE) at Johns Hopkins University. All code files and results are publicly available at https://github.com/RajDandekar/Reopening ImpactSimulator US States.

## 8 ACKNOWLEDGEMENTS

This effort was partially funded by the Intelligence Advanced Reseach Projects Activity (IARPA). We are grateful to Haluk Akay, Hyungseok Kim and Wujie Wang for helpful discussions and suggestions.

## 9 AUTHOR CONTRIBUTIONS

R.D. and G.B. designed the research. C.R. and R.D. designed the model framework. R.D. and E.W. applied the model to all the states considered. R.D., C.R., E.W. and G.B. wrote the study.

## 10 RESOURCE AVAILABILITY

**Lead Contact:** Raj Dandekar, MIT.

